# Racial/Ethnic Disparities in the Observed COVID-19 Case Fatality Rate Among the U.S. Population

**DOI:** 10.1101/2022.03.01.22271708

**Authors:** L. Philip Schumm, Mihai C. Giurcanu, Kenneth J. Locey, Jean Czerlinski Ortega, Zhenyu Zhang, Robert L. Grossman

## Abstract

**Purpose:** During the initial 12 months of the pandemic, racial and ethnic disparities in COVID-19 death rates received considerable attention but it has been unclear whether disparities in death rates were due to disparities in case fatality rates (CFRs), incidence rates or both. We examined differences in observed COVID-19 case fatality rates (CFRs) between U.S. Whites, Blacks/African Americans and Latinx during this period.

**Methods:** Using data from the COVID Tracking Project (CTP) and the CDC’s COVID-19 Case Surveillance Public Use dataset, we calculated CFR ratios comparing minority groups to Whites, both overall and separately by age group. We also used a model of monthly COVID-19 deaths to estimate CFR ratios, adjusting for age, gender, and differences across states and time.

**Results:** Overall Blacks and Latinx had lower CFRs than Whites. However, when adjusting for age, Blacks and Latinx had higher CFRs than Whites among those younger than 65. CFRs varied substantially across states and time.

**Conclusions:** Disparities in COVID-19 case fatality among U.S. Blacks and Latinx under age 65 were evident during the first year of the pandemic. Understanding racial/ethnic differences in COVID-19 CFRs is challenging due to limitations in available data.

## Introduction

In the U.S., minority groups including Blacks/African Americans and Latinx have experienced higher rates of COVID-19 infection and mortality than Whites, as well as higher rates of excess mortality associated with COVID-19.^1–3^ Assessing differences in outcomes among those infected is challenging at the national level due to limitations in available data. Studies among subpopulations have often found little or no evidence of racial/ethnic disparities in case fatality. Using data from the Michigan Disease Surveillance System (MDSS) from March 8 through July 5, 2020, Zelner et al. found that Blacks had substantially higher rates of COVID-19 infection and mortality. However, the case fatality rate among Blacks was only modestly higher than that of Whites, suggesting that differences in COVID-19 mortality were driven by differences in the rate of infection.^4^ Similarly, among individuals receiving care in the US Department of Veterans Affairs tested for COVID-19 between February 8 and July 22, 2020, Rentsch et al. found that while Blacks and Hispanics were more likely to test positive than Whites, once infected their 30-day mortality was less and, after adjusting for age, sex and rural/urban residence, was equivalent to that of Whites.^5^

Several studies have examined racial/ethnic differences in hospitalization and/or case fatality rates among health care systems, where access to electronic health records permits adjusting for demographic and clinical covariates. Azar et al. examined COVID-19 positive patients within Sutter Health in northern California from January 1 through April 8, 2020, and found that African Americans had 2.7 times the odds of hospitalization compared to Whites despite adjusting for age, sex, comorbidities and income.^6^ Similarly, Price-Haywood et al. studied COVID-19 positive patients within Ochsner Health in Louisiana and found that although Blacks had roughly twice the odds of hospitalization than Whites, their risk of in-hospital death was similar.^7^ Adjusting for demographics, income, comorbidities and clinical characteristics upon admission had only a modest impact on the estimated difference in mortality. Finally, Ogedegbe et al. examined patients within New York University’s Langone Health system who were tested for COVID-19 between March 1 and April 8, 2020.^8^ In contrast to the previous two studies, the odds of hospitalization among those who tested positive were similar among Blacks, Hispanics and Whites. However, among those hospitalized, Black and Hispanic patients were less likely than White patients to die despite adjusting for demographic characteristics, comorbidities and insurance.

Griffith et al. discuss the potential of collider bias to distort understanding of COVID-19 disease risk and severity.^9^ One potential source of collider bias is restricting analyses to a non-representative subset of the population, such as those hospitalized for an active COVID-19 infection. For example, it is possible that a tendency among Blacks/African Americans to seek care later in the acute setting—perhaps due to structural inequities, biases on the part of providers and prior negative experiences with health care—may partly account for the higher rates of admission observed in some studies.^6,10^ If true, focusing on in-hospital mortality could reduce the apparent difference in case fatality between Blacks and Whites due to a larger proportion of Black individuals with COVID-19 being hospitalized. More generally, while studies of selected subgroups can benefit from higher quality data and can answer questions of local interest, it remains important to study COVID-19 risk and severity at the population level.

The purpose of this study was to examine racial/ethnic differences in COVID-19 case fatality rates (CFRs) at the U.S. national level during the first year of the pandemic. We used data from The Atlantic’s COVID Tracking Project (CTP), whose Racial Data Tracker was widely regarded as the most complete source of information on race/ethnicity of COVID-19 cases and deaths during this period. We performed a parallel analysis using the Centers for Disease Control and Prevention (CDC) COVID-19 Case Surveillance Public Use data—an independently compiled and regularly updated individual-level data source. Although less complete than the CTP data, the CDC data contain information on age. Adjusting for age is critical to get an accurate understanding of differences in COVID-19 CFR.^11^ We focused on the two largest minority groups, Blacks/African Americans and Latinx, and their comparison to Whites, since these categories are reported most completely by a large number of states and territories, and permit approximate comparability between the two datasets.

## Methods

### Data

#### The COVID Tracking Project (CTP)

The COVID Tracking Project (CTP) was a volunteer-run effort started in March 2020 to compile and publish nationwide data on the COVID-19 pandemic that were not available elsewhere. Volunteers manually harvested and curated data from individual states’ public health websites on a daily basis. The CTP ceased operations in March 2021 due to improvements in the quality and timeliness of information being provided by the CDC and HHS. During its operation, the CTP was considered an authoritative source of COVID-19 data, and their data have been used in over 1,000 academic articles. CTP datasets remain available on their website, and may be used under the Creative Commons CC BY 4.0 license.^12^

The COVID Racial Data Tracker, part of the CTP, collected state-level data on cases and deaths separately by race/ethnicity for the purpose of examining the disproportionate impact of the pandemic on minority communities. The dataset contains twice weekly counts of the cumulative number of cases and deaths for each state for which information was available. Separate counts by racial group (White, Black, Latinx, Asian, American Indian/Alaskan Native, Native Hawaiian/Other Pacific Islander, Multiple, or Other) and ethnicity (Hispanic or Non-Hispanic) are also provided. Because information on the joint distribution of race and ethnicity was not provided, we utilized information on racial group only (in many cases, the counts for Latinx were the same as for Hispanic). Information on age is not available.

Our analyses of CTP data used cumulative counts as of February 28, 2021—the last full month for which data were provided. This allowed us to construct a corresponding dataset from the CDC data which are recorded on a monthly basis.

#### CDC COVID-19 Case Surveillance Public Use Data

This dataset contains individual-level data on all cases reported to the CDC.^13^ The public use dataset contains 19 data elements for each case, including the state and month in which the case was reported, whether the individual died as a result of COVID-19, and individual-level characteristics such as sex (Male or Female), age group (0–17, 18–49, 50–64 and 65+), race (White, Black, Asian, American Indian/Alaskan Native, Native Hawaiian/Other Pacific Islander, Multiple/Other) and ethnicity (Hispanic or Non-Hispanic). We define the three racial/ethnic groups for our analysis as follows: White non-Hispanic, Black, and Hispanic non-Black. For comparability with the CTP dataset, we use all cases reported through February 2021.

### Statistical Analysis

Case fatality rates (CFR) were computed by dividing the total number of deaths by the total number of cases reported as of February 28, 2021. Monthly rates for the CTP data were computed by dividing the total number of new deaths reported in that month by the total number of new cases. While some deaths reported in a month corresponded to cases reported from previous months, sensitivity analyses including cases from the 2–3 previous months yielded similar results. Monthly rates for the CDC data were computed as the proportion of cases reported in that month that resulted in death; this is therefore a leading indicator since some of those deaths occurred in subsequent months. The following states or territories were excluded from all analyses of the CDC data: nine states (AK, DE, HI, MO, NE, SD, TX, VI, WV) reported no deaths, with survival status missing for all or nearly all cases; three states reported only deaths (WA), nearly all deaths (IL) or half as many deaths as cases (RI), also with survival status missing for a large fraction of cases; and one state (GA) and Puerto Rico were outliers in a log-log plot of deaths versus all cases (including those with missing survival status). These exclusions reduced our analyses of CDC data to 38 states. When computing the CFR for a given racial/ethnic group, only those states reporting both cases and deaths for that group were included.

Since the distribution of racial/ethnic groups differs across states, state-specific differences in the CFR may bias comparisons between groups. Thus, we calculated CFR ratios comparing each minority group to Whites separately by state and estimated an overall CFR ratio using a random effects model:

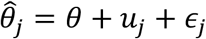

Where 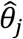 is the natural log of the CFR ratio for state *j*, the *u*_*j*_ ~ *N*(0, *τ*^2^) represent between-state differences in the log CFR ratio, and the 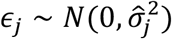 represent sampling variability. The between-state variance (*τ*^2^) was estimated using the DerSimonian–Laird method,^14^ and an estimate of the overall CFR (*θ*) was obtained as a weighted sum of the 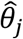 this estimate was then exponentiated to obtain a CFR ratio, together with the endpoints of the corresponding 95% Wald confidence interval. The *I*^2^ measure of heterogeneity (representing the percentage of variability in the 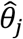 due to between-state differences) is reported.^15^

To estimate CFR ratios adjusting for differences in the CFR across time, gender, age group and state, we used the CDC data to compute the number of cases (*c*_*ijk*_) and deaths (*d*_*ijk*_) for each demographic subgroup *i* (based on the Cartesian product of age group, gender and race/ethnicity), state *j*, and month *k*, and fit the following mixed-effects Poisson model to the data for Whites, Blacks and Latinx age 18 and older:^16^

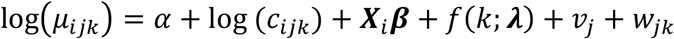

where *µ*_*ijk*_ is the expected number of deaths, ***X***_*i*_ is a vector of discrete covariates describing the *i*th group including age group, gender, race/ethnicity and an interaction between age group and race/ethnicity, and *f*(*k; λ*) is a restricted cubic spline function of *k* depending on coefficients *λ* that captures the change over time (we used 5 knots based on Harrell’s recommended percentiles).^17^ The term *υ*_*ijk*_ is a random effect capturing differences between states, and the term ***w***_*jk*_ is a random effect, nested within state, capturing remaining differences across time within state. Variance estimates were obtained using the clustered form of the robust (sandwich) variance estimator with clustering at the state level.^18^ Estimated coefficients (***β***) were exponentiated to obtain CFR ratios, together with the endpoints of their corresponding 95% Wald confidence intervals. Estimates of the *υ*_*j*_ were obtained using empirical Bayes means and plotted on a map.

All analyses were performed using Stata version 17.0, Python, pandas and geopandas (for data management) and Altair (for plotting).^19^ Data and code are available on GitHub (https://github.com/pschumm/covid-modeling).

## Results

The CTP dataset includes 28.4 million cases and 512,627 deaths as of February 28, 2021, corresponding to an overall CFR of 1.8% (Table 1). The CDC dataset contains 22.1 million cases over the same period, however survival status is missing for nearly half (10.4 million). Among the other 11.7 million cases, there were 263,398 deaths corresponding to an overall CFR of 2.3%. Removing those states and territories with problematic data (see Methods) left 235,635 deaths with an overall CFR of 2.0%. Both datasets show a similar pattern in the CFR over time, with the CDC curves running ahead of the CTP curves by approximately 1–2 months, as expected (Figure 1). Based on the CDC data, the CFR decreased from the start of the pandemic through September/October 2020. This decrease was followed by an increase and a subsequent decline, consistent with the estimate of *f*(.) from the mixed-effects model when fit to the CDC data below (Figure 2).

**Table 1.** Case fatality rates (CFR) for COVID Tracking Project (CTP) and CDC public use datasets through February 2021

|  | Deaths | Cases | CFR (%) |
| --- | --- | --- | --- |
| *CTP* |  |  |  |
| Overall | 512,627 | 28,443,555 | 1.8 |
| Race/Ethnicity |  |  |  |
| White<sup>a</sup> | 268,373 | 10,309,363 | 2.6 |
| Black/AA<sup>b</sup> | 61,989 | 2,359,473 | 2.6 |
| Latinx<sup>c</sup> | 71,844 | 3,666,785 | 2.0 |
| Other | 39,403 | 2,539,435 | 1.6 |
| Missing | 34,715 | 9,510,434 | 0.4 |
| *CDC* |  |  |  |
| Overall | 263,398 | 11,700,999<sup>d</sup> | 2.3 |
| Post-filtering<sup>e</sup> | 235,635 | 11,661,626 | 2.0 |
| Age group<sup>e</sup> |  |  |  |
| < 18 | 0 | 1,327,892 | 0.0 |
| 18–49 | 3,728 | 6,338,709 | 0.1 |
| 50–64 | 18,689 | 2,316,705 | 0.8 |
| 65+ | 213,077 | 1,580,396 | 13.5 |
| Missing | 141 | 97,924 | 0.1 |
| Race/Ethnicity<sup>e</sup> |  |  |  |
| White | 132,822 | 3,955,817 | 3.4 |
| Black/AA | 22,735 | 861,823 | 2.6 |
| Latinx | 26,191 | 1,158,313 | 2.3 |
| Other | 11,074 | 854,367 | 1.3 |
| Missing | 42,813 | 4,831,306 | 0.9 |

<sup>a</sup>Excludes the following states/territories for which counts of cases and/or deaths are not available: AS, MP, ND, NY, PR and VI.
<sup>b</sup>Excludes the following states/territories for which counts of cases and/or deaths are not available: AS, GU, MP, ND, NY, PR and VI.
<sup>c</sup>Excludes 33 states/territories for which counts of cases and/or deaths are not available (23 states remaining).
<sup>d</sup>Excludes 10,371,233 cases for which information on survival is not available.
<sup>e</sup>Includes data from 38 states that remained after QC filtering.

**Figure 1.**
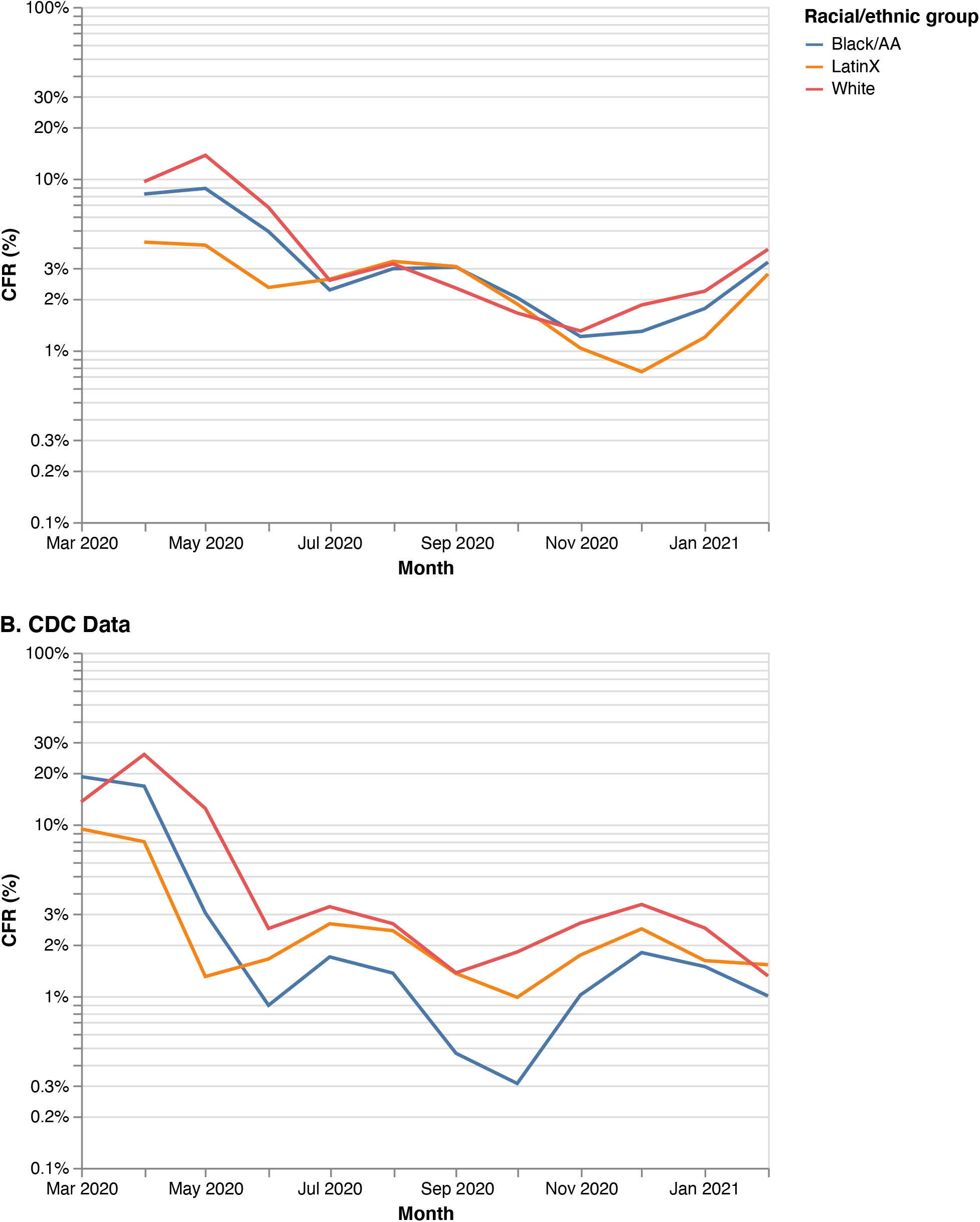
CFR by month and racial/ethnic group, CTP and CDC datasets.

**Figure 2.**
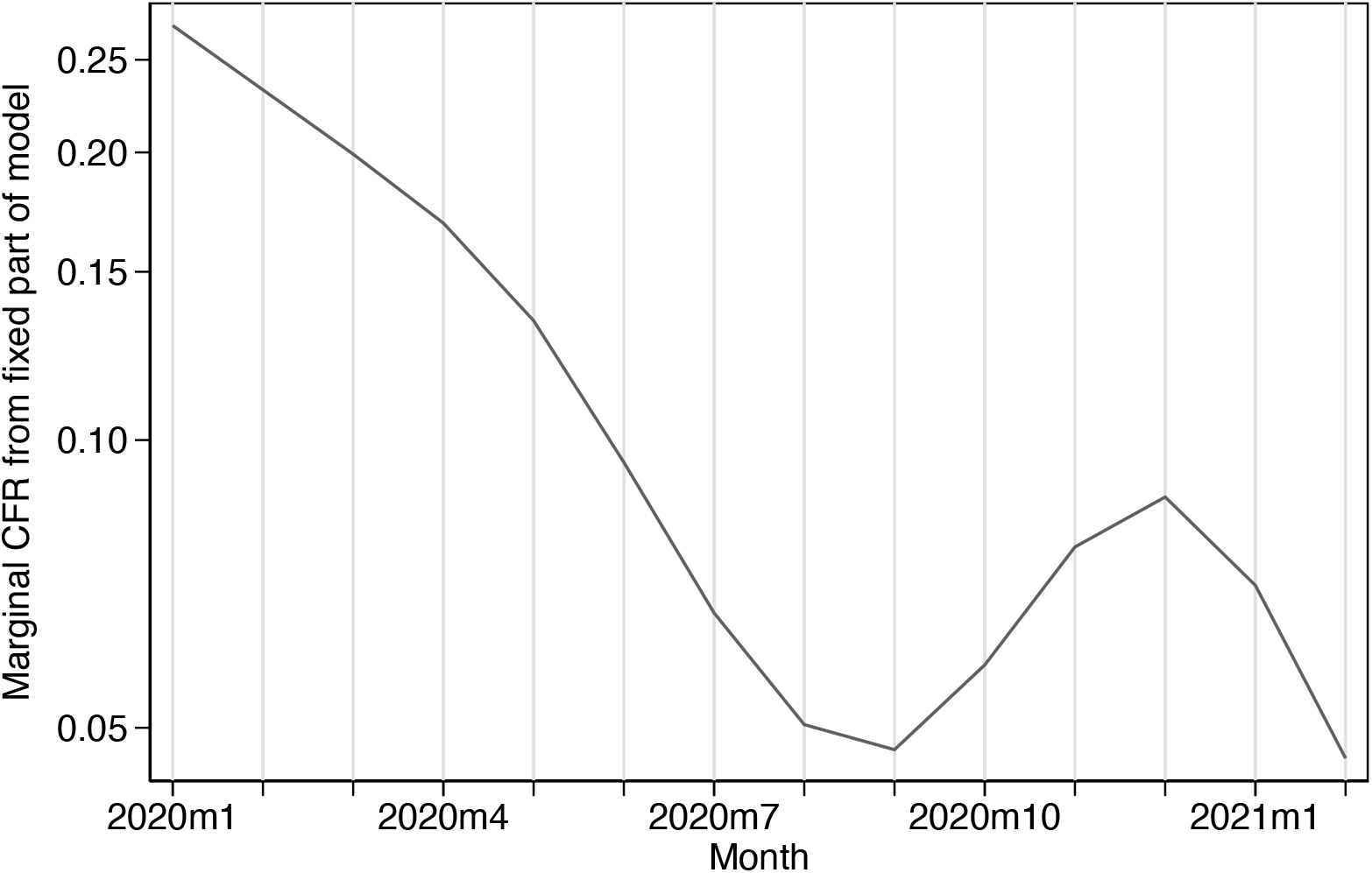
Estimated overall time trend in CFR from mixed-effects model fit to CDC data, parameterized using restricted cubic spline with 5 knots.

The CFR was substantially higher for those 65 and older (13.5%) as compared to those 50–64 (0.8%) and 18–49 (0.1%) (Table 1). No deaths were reported in the CDC dataset for individuals under 18 years of age. In the CDC dataset, the CFRs for Black and Latinx were lower than for Whites throughout the entire year (Figure 1). In the CTP dataset, CFRs for the three racial/ethnic groups were similar from July 2020 through November 2020, though minorities still had lower CFRs than Whites before and after this period. Table 2 (Panel A) shows estimated CFR ratios comparing the two minority groups to Whites, after adjusting for differences in CFR across states. In the CTP dataset, the CFR ratio comparing Blacks to Whites was 0.8 (95% CI [0.7, 0.9]) and comparing Latinx to Whites was 0.4 (95% CI [0.3, 0.5]); the corresponding CFR ratios for the CDC dataset were even lower at 0.5 (95% CI [0.4, 0.6]) and 0.2 (95% CI [0.2, 0.3]), respectively.

**Table 2.** Case fatality rate (CFR) ratios comparing minority groups to Whites, estimated from state-specific log CFR ratios using random-effects models (CTP and CDC) and based on a mixed-effects Poisson regression model fit to individual-level data (CDC only)

|  | *Black/AA compared to White* |  |  | *Latinx compared to White* |  |  |
| --- | --- | --- | --- | --- | --- | --- |
|  | No. states | Rate ratio | 95% CI | No. states | Rate ratio | 95% CI |
| *A. Random-effects models fit to state-specific log CFR ratios* |  |  |  |  |  |  |
| *CTP* |  |  |  |  |  |  |
| Overall | 49 | 0.8 | (0.7, 0.9) | 23 | 0.4 | (0.3, 0.5) |
| *CDC* |  |  |  |  |  |  |
| Overall | 26 | 0.5 | (0.4, 0.6) | 14 | 0.2 | (0.2, 0.3) |
| Age group |  |  |  |  |  |  |
| 18–49 | 2 (CA & NY) | 3.9 | (0.3, 44.5) | 3 (AZ, CA & NY) | 9.8 | (4.4, 21.9) |
| 50–64 | 12 | 2.1 | (1.2, 3.6) | 9 | 3.0 | (2.2, 3.9) |
| 65+ | 26 | 0.9 | (0.8, 1.1) | 13 | 0.9 | (0.8, 1.2) |
| *B. Mixed-effects Poisson regression model* |  |  |  |  |  |  |
| *CDC* |  |  |  |  |  |  |
| Age group |  |  |  |  |  |  |
| 18–49 |  | 6.0 | (3.4, 10.8) |  | 4.1 | (1.9, 8.7) |
| 50–64 |  | 2.7 | (1.8, 4.1) |  | 2.4 | (1.5, 3.8) |
| 65+ |  | 1.0 | (0.8, 1.1) |  | 1.2 | (1.0, 1.4) |

The proportion of cases under age 65 was higher among Blacks (89%) and Latinx (93%) than among Whites (80%). These differences were even larger for the proportion of cases under age 50 (69% for Blacks and 78% for Latinx as compared to 58% for Whites). When stratifying by age group, the state-specific CFRs—for states reporting any deaths for a particular age by racial/ethnic group—among those aged 18–49 and 50–64 were higher, on average, among Blacks and Latinx than among Whites (Figure 3). The CFR ratios comparing Blacks to Whites were 3.9 for ages 18–49, 2.1 for ages 50–64 and 0.9 for ages 65 and older; corresponding CFR ratios comparing Latinx to Whites were 9.8, 3.0 and 0.9, respectively (Table 2, Panel A).

**Figure 3.**
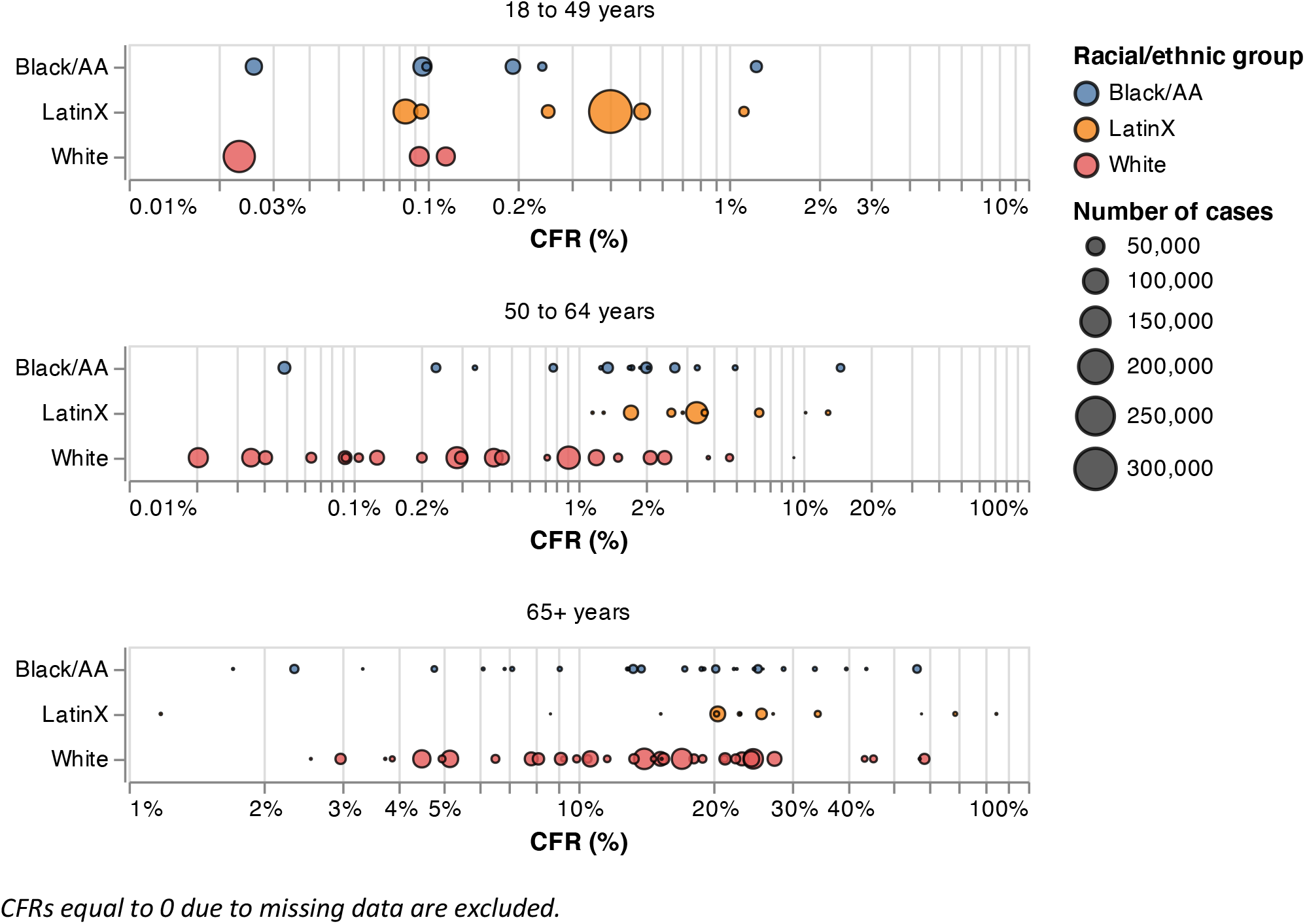
State-specific CFRs, separately by age group and racial/ethnic group (CDC data)

Results from the mixed-effects model fit to deaths reported in the CDC data are shown in Table 2 (Panel B). Consistent with the CFR ratios reported above, these results demonstrated higher CFRs for minority groups among those under age 65. Specifically, estimated CFR ratios comparing Blacks to Whites were 6.0 (95% CI [3.4, 10.8]) for ages 18–49, 2.7 for ages 50–64 (95% CI [1.8, 4.1]) and 1.0 (95% CI [0.8, 1.1]) for ages 65 and older. The corresponding estimated CFR ratios comparing Latinx to Whites were 4.1 (95% CI [1.9, 8.7]), 2.4 (95% CI [1.5, 3.8]) and 1.2 (95% CI [1.0, 1.4]), respectively. The CFR ratio comparing women to men (not shown) was 0.76 (95% CI [0.72, 0.81).

We observed considerable variation between states for both datasets, both in the CFRs and in the CFR ratios comparing racial/ethnic groups (Figure 3). The value of *I*^2^ was over 95% in all cases. The estimated variance of the *υ*_*j*_ was 0.31, corresponding to an increase in the CFR of 75% for a state one standard deviation above the mean. Similarly, the estimated variance of the ***w***_*jk*_ was 0.26, corresponding to a 67% increase in the CFR for a month one standard deviation above the mean; this variability within state over time is above and beyond that already accounted for by the estimated overall time trend 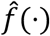 (Figure 2). Figure 4 plots estimates of the *υ*_*j*_ for all 38 states included in the model.

**Figure 4.**
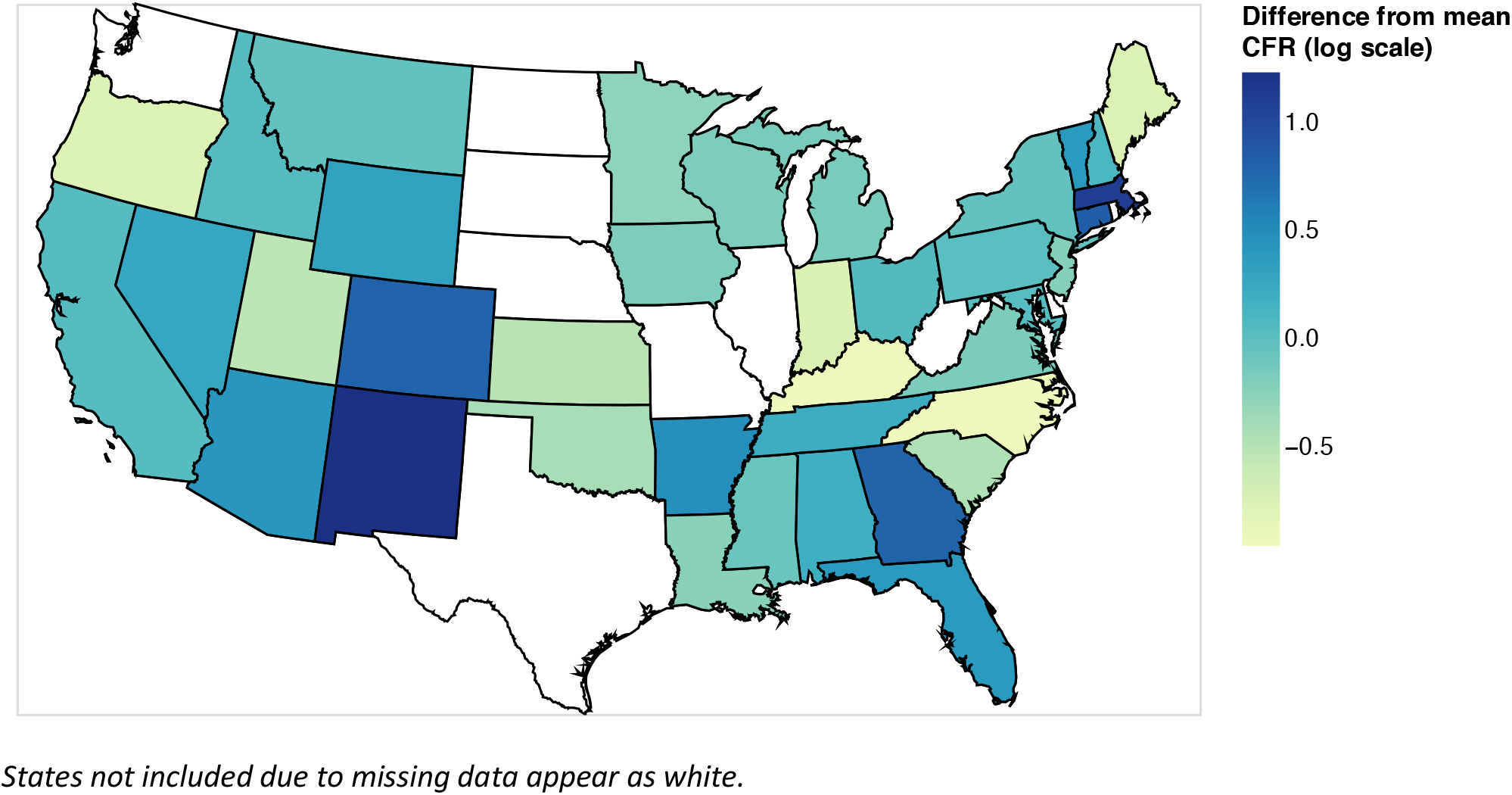
Estimated differences in CFR across states (CDC data)

## Discussion

While the overall CFR was lower among Blacks and Latinx than among Whites, the CFRs for Blacks and Latinx within age category were higher among those under age 65. This reversal in the direction of the association when stratifying by subgroup is a partial example of what is known as Simpson’s Paradox.^20^ Although a full reversal of the association was not evident among those 65 or older (with the exception of the mixed-model estimated CFR ratio of 1.2 comparing Latinx to White), it is possible that further adjustment for age within this oldest subgroup would yield a reversal here too. Thus, these data reinforce the point made by Green et al. that COVID-19 CFRs that are not age-specific “may hide more than they reveal.”

We also found evidence of an interaction between age and race/ethnicity such that Black and Latinx disparities in COVID-19 CFR were largest among the youngest ages. To our knowledge, this finding has not been previously reported. This finding deserves further study as it could have substantial implications for health policy—especially for a disease where public attention concerning mortality, at least during the early stages of the pandemic, was primarily focused among the oldest ages.

Our results reflect differences in the *observed* CFR; that is, the number of deaths divided by the number of reported cases. For COVID-19, this differs substantially from the true, underlying CFR since many infected individuals, especially those who were asymptomatic, were never tested. Moreover, some individuals who tested positive may not appear in the datasets used here. Thus, we expect that much of the change over time, as well as of the substantial variation across states, reflects variation in the rates of testing and reporting. Several studies have found higher testing rates among Blacks and Hispanics than among Whites, including a study of roughly 50 million patients in the Epic health record system.^5,21–23^ Higher testing rates among minorities would be expected *ceteris paribus* to reduce their CFR relative to Whites. Furthermore, by adjusting for differences across time and between states, our results should be robust to confounding due to differences in the racial mix of cases over time or in the distribution of minorities across states.

The CDC dataset contains only 78% of the cases in the CTP dataset, and of these, information on survival is missing for 47% of cases. In addition, among the cases with survival status for the 38 states included in the analysis, race/ethnicity was missing for 41%. Despite these limitations, analysis of this dataset correctly revealed the improvements in COVID-19 survival over the first 6–8 months of the pandemic as well as the established lower risk of death for women relative to men.^24–26^ While the overall CFR ratios comparing minorities to Whites were smaller than for the CTP dataset, the basic conclusion is the same.

Finally, though we found evidence of racial/ethnic disparities in CFR among those under age 65, our study was also intended to highlight the limitations in data available on race/ethnicity of COVID-19 cases and deaths at the national level. Specifically, using information on the joint distribution of race/ethnicity and age (as available in the CDC dataset only) reveals an entirely different understanding than when not using it. Chowkwanyun and Reed argue convincingly that while identifying racial/ethnic disparities in COVID-19 is important, a lack of context as provided by richer data “can perpetuate harmful myths and misunderstandings that actually undermine the goal of eliminating health inequities.”^27^ Acquiring such data from representative samples of the national population is sorely needed.

## Data Availability

All data files and code used in the analyses reported here are available online at https://github.com/pschumm/covid-modeling.

